# Disparities in Age-Specific Morbidity and Mortality from SARS-CoV-2 in China and the Republic of Korea

**DOI:** 10.1101/2020.03.24.20042598

**Authors:** Joseph P. Dudley, Nam Taek Lee

## Abstract

We analyzed age-specific and sex-specific morbidity and mortality data from SARS-COV-2 pandemic in China and Republic of Korea (ROK). Data from China exhibit a standard Gaussian distribution with peak morbidity in the 50-59 years cohort, while the ROK data have a bimodal distribution with highest morbidity in the 20-29 years cohort.

## INTRODUCTION

There are many uncertainties regarding the clinical epidemiology of the SARS-CoV-2 virus that is currently spreading worldwide in a global pandemic whose ultimate impacts are still uncertain, but which appears based on observed impacts to have the potential to overwhelm the medical surveillance and medical treatment infrastructure of even the world’s most affluent countries. There is a need to gain greater understanding of the highest risk populations for infection and serious disease from the SARS-CoV-2 virus to support the development and implementation of effective public health surveillance and mitigation efforts, and minimize the adverse effects of the current COVID-19 Pandemic in countries worldwide [1].

## METHODS

We analysed published data on age and sex of SARS-CoV-2 cases published by the Korea Centers for Disease Control & Prevention (KCDC) [2], and the China Centers for Disease Control [3] to compare the age-specific rates of morbidity and mortality from the recent COVID-19 pandemic in China and the Republic of Korea, and gain insights into the potential for variation in public health impacts of the pandemic in other countries worldwide.

## RESULTS

We identified major disparities in the age-specific and sex-specific rates of morbidity and mortality from SARS-CoV-2 in China and the ROK.

Data from China (n = 44,672 as of 11 February 2020) exhibit a typical symmetrical Gaussian configuration, with peak morbidity among individuals in the 50-59 year age cohort (Figure 1). The sex-specific morbidity ratio among confirmed cases was near parity, with a slight male bias (male 51%; female 49%). There was a high degree of difference in sex-specific case fatality rates, with fatality rates much higher among males (2.8% CFR) than females (1.7% CFR).

**FIGURE 1.**
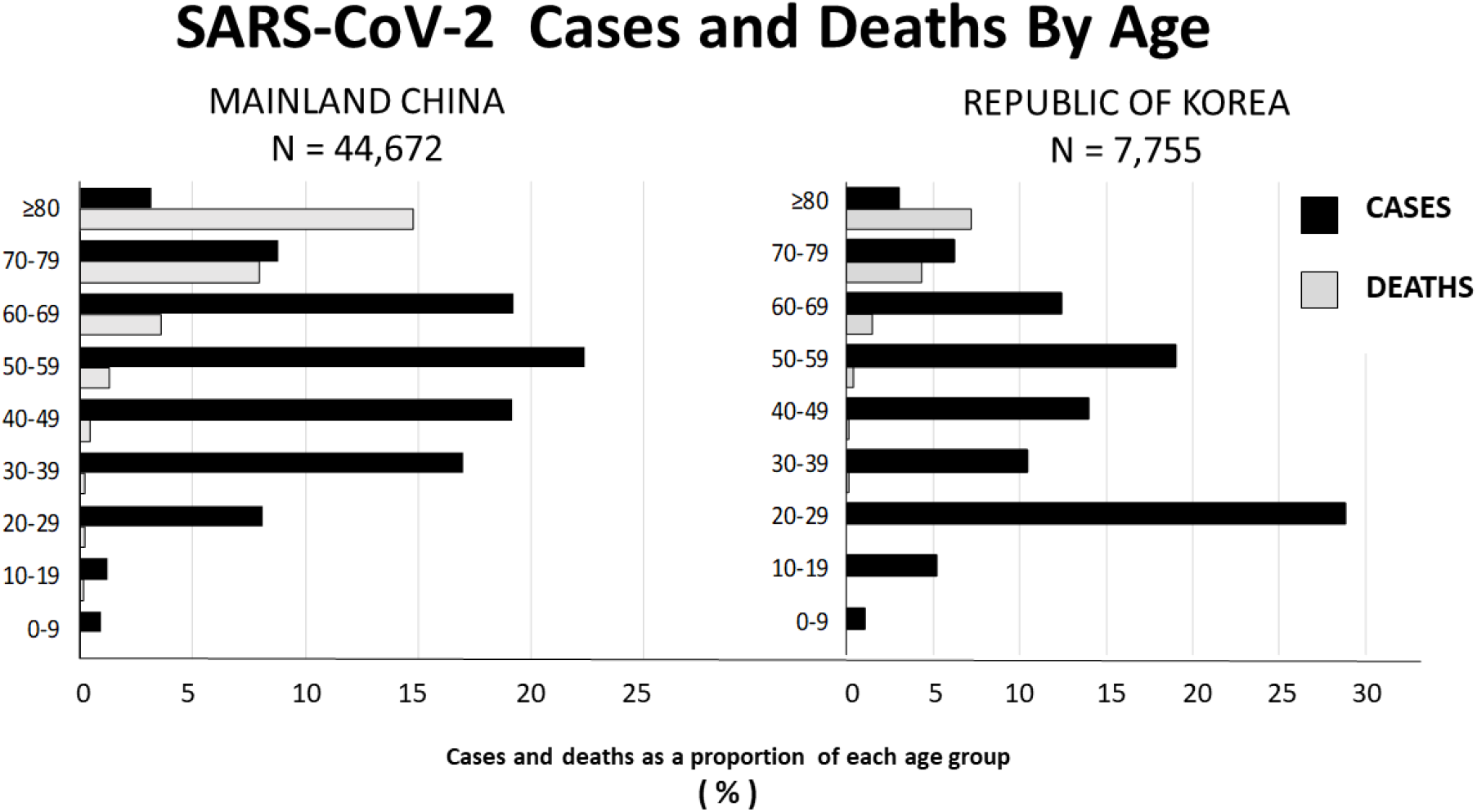
Comparison of Age-Specific Morbidity and Mortality Rates Among Reported Laboratory Confirmed Cases from China and Republic of Korea

Data from ROK (n = 7,755 as of 11 March 2020) exhibited a skewed bimodal distribution with the highest rate of confirmed SARS-CoV-2 infections among individuals in the 20-29 year age cohort (30%), and a second peak in the 50-59 year age cohort (19%) (Figure 1). There is a high degree of difference in the sex-specific rates of infection from SARS-CoV-2 in the ROK, with female cases (62%) outnumbering males by a factor of nearly 2:1 ratio (females: 62%; males: 38%). The data on fatalities from confirmed SARS-CoV-2 infections in the ROK exhibit a reversed bias ratio to that among cases, with the case fatality rate among males more than twice as high as that among females (CFR: males 1.19%; females 0.52%).

## DISCUSSION

Investigations of age-specific and sex-related differences in morbidity and mortality from emerging diseases can provide important tools for identifying populations at highest risk for surveillance, monitoring and intensive medical interventions, and provide insights into geographic, genetic or cultural factors that may influence the spread of pandemic disease viruses such SARS-CoV-2, within and among different countries and continents [4-5].

The age-specific and sex-specific profiles of the current SARS-CoV-2 pandemic in the ROK are quite different from those from the MERS-CoV epidemic that occurred during May-June 2015, which was the largest outbreak of the MERS-CoV virus outside of the Middle East region and involved a total of 186 confirmed cases, including 38 fatalities (20% CFR). The epidemic lasted for 2 months and the government quarantined 16,993 individuals for 14 days to control the epidemic. The 2015 MERS-CoV epidemic in ROK was almost entirely nosocomial, through intra-hospital and hospital-to-hospital nosocomial transmission, rather than community transmission. Only one of the 186 confirmed cases from the 2015 MERS-CoV epidemic occurred through community transmission [6].

The age-specific morbidity and mortality rates from the 2015 MERS-CoV epidemic in ROK are different in certain respects from those of the SARS-CoV-2 pandemic in China and the ROK. The general shape of the epidemic curve from the 2015 Korea MERS-CoV outbreak was a standard unimodal Gaussian curve with peak morbidity in the 50-59 year age cohort, similar to that of the SARS-CoV-2 pandemic for China shown in Figure 1. The sex ratio among confirmed cases from the 2015 Korea MERS-CoV epidemic in the ROK was male biased, with a 60:40 ratio of males to females, the reverse of that observed in the current SARS-CoV-2 pandemic, which has a 40:60 male female ratio among confirmed cases reported as of 11 Mar 2020 [6].

Although nosocomial transmission has been an important factor in the current SARS-CoV-2 pandemic in both China and the ROK, community transmission has been the predominant driving factor for the current SARS-CoV-2 pandemic in ROK. Improvements in hospital infection control policies and practices, coupled with the development and implementation of more sophisticated infectious disease surveillance and response training programs, are believed to be important factors in the apparent relatively low rate of nosocomial transmission in the ROK during the current pandemic.

The available data indicate that the epidemiological characteristics of SARS-CoV-2 infections appear to distinctly different from those of typical human coronaviruses and the recently-emerged zoonotic coronaviruses (SARS-CoV, MERS-CoV), zoonotic influenza viruses (e.g., H5N1, H7N9, H792), seasonal influenza viruses, and recent pandemic influenza viruses [7]. While many patients developed influenza-like illness that progresses to pneumonia or acute respiratory distress syndrome (ARDS) resulting in hospitalization and/or death, a large spectrum of clinical presentations have been documented including a growing number of reported cases with mild symptoms and asymptomatic infections [8]. Asymptomatic infection has been reported from adults and children, but the proportion of truly asymptomatic infections remains uncertain, and may be higher than documented from currently available data [9-10]. A recent report documented RT-PCR detection of SARS-CoV-2 in fecal samples from an asymptomatic child, which persisted for 26 days following the estimated date of exposure [11].

The most commonly reported symptoms in order of relative frequency include fever and dry cough (>70%) and fatigue (>35%), dyspnea (<20%), sore throat (<15%), headache (<15%), myalgia or arthralgia (<15%), chills (<15%), nausea or vomiting (5%), nasal congestion (<5%), diarrhea (<4%), and conjunctivitis (<1%). People with symptomatic COVID-19 infections usually become symptomatic within 1-2 weeks following infection (mean incubation period 5-6 days, range 1-14 days). The majority of people infected with COVID-19 virus have mild disease and recover. Approximately 15% of laboratory confirmed symptomatic patients develop severe disease with pneumonia and/or dyspnea, of whom about 7% progress to respiratory failure, septic shock, or multiple organ failure). Individuals at highest risk for severe disease and death include people aged over 60 years and individuals with chronic disease. Symptomatic disease appears to be relatively rare and typically mild in children and adolescents [7].

We hypothesize that differences in public health intervention practices and age-related sociocultural factors may be significant factors mediating the observed marked disparities in the age-specific and sex-specific rates of infection from confirmed cases in China and Korea. The most striking anomaly in the ROK data appears to be the relatively high proportion of cases among the 20-29 year age group (29% of all cases), which may be attributable in part to lower rates of compliance among individuals in this age group with social distancing and self-quarantine recommendations issued by Korean health authorities. Religious affiliations and practices have been an important factor for the SARS-CoV-2 pandemic in the ROK, because approximately 61% of all confirmed cases as of 11 Mar 2020 have epidemiological links to the Shincheonji religious community, a Korean-based sect with satellite churches in China and at least 24 other countries, with some 200,000 members in ROK (∼0.3% of the ROK population of 51.5 million persons) [12]. Lack of adherence to social-distancing and self-quarantine recommendations appears to be a key factor in the high ratio of confirmed SARS-CoV-2 infected individuals associated with the Shincheonji religious community also. Observational data collected throughout the course of the COVID-19 pandemic in the ROK indicates much lower rates of compliance with ROK government social distancing recommendations by individuals in the young adults and teenagers in the 15-29 year age cohorts, which could help explain the relatively higher rates of infection among the 10-19 and 20-29 year age cohorts in the ROK data.

## CONCLUSIONS

The reported data on confirmed cases and fatalities from SARS-CoV-2 indicates highly significant differences in the age-specific and sex-specific rates of morbidity and mortality from the COVID-19 Pandemic in the Republic of Korea (ROK) and China. Closer study of the factors underlying the initiation and early proliferation of the SARS-CoV-2 epidemics in China and the ROK may provide valuable insights in the development of more effective strategies for predicting and mitigating the spread of the COVID-19 pandemic in other countries worldwide.

Improvements in hospital infection control policies and practices, and the development and implementation of cross-sectoral civilian and military infectious disease outbreak surveillance and response training programs by the ROK government in coordination with international partner organizations such as the WHO and US Centers for Disease Control and Prevention are believed to be important factors in the apparent relatively low rate of nosocomial transmission in the ROK during the current COVID-19 pandemic, and the effectiveness of the ongoing public health surveillance and monitoring programs.

The available epidemiological and observational data from the ROK suggests that reduced rates of compliance with social distancing and self-quarantine recommendations among different sectors of the population – especially the younger adult and juvenile age cohorts -- may have a significant impact on the age-specific rates of morbidity and mortality within the population as a whole.

## Data Availability

data are available from from source references and urls provided in REFERENCES section, and below

https://www.cdc.go.kr/board/board.es?mid=a30402000000&bid=0030&act=view&list_no=366523&tag=&nPage=1

## Acknowledgments

Ian M. Mackay provided review and comments on a preliminary draft, source reference citation and tabular summary data on reported cases from China. Donald J. Kosiac provided review and comments on draft manuscript. No external, corporate or institutional funding support was received for this work.

